# Investigating the Determinants of COVID-19 Vaccine Uptake and Attitudes in Iraq: A Study Unveiling the Negative Impact of Misinformation and Vaccine Conspiracy

**DOI:** 10.1101/2023.12.06.23299628

**Authors:** Malik Sallam, Nariman Kareem, Mohammed Alkurtas

## Abstract

Vaccine hesitancy is a major barrier challenging the control of infectious diseases. Previous studies demonstrated high rates of COVID-19 vaccine hesitancy in the Middle East. The current study aimed to investigate the attitudes towards COVID-19 vaccination and COVID-19 vaccine uptake among the adult population in Iraq. This cross-sectional self-administered survey-based study was conducted in August–September 2022. Recruitment of possible participants was done using chain-referral sampling. The survey instrument assessed participants’ demographics, attitudes toward COVID-19 vaccination, beliefs in COVID-19 misinformation, vaccine conspiracy beliefs, and sources of information regarding the vaccine. The study sample comprised a total of 2544 individuals, with the majority reporting the uptake of at least one dose of COVID-19 vaccination (n=2226, 87.5%). Positive attitudes towards COVID-19 vaccination were expressed by the majority of participants (n=1966, 77.3%), while neutral attitudes were expressed by 345 participants (13.6%), and negative attitudes were expressed by 233 participants (9.2%). Strong, moderate, slight, and absence of COVID-19 misinformation were expressed by 12.4%, 22.6%, 36.2%, and 28.7% participants, respectively. The majority of participants showed a neutral attitude towards COVID-19 vaccine conspiracies (n=1464, 57.5%), while 607 participants embraced these conspiracies (23.9%), and 473 disagreed with such beliefs (18.6%). In the multivariate analysis, factors associated with positive attitudes towards COVID-19 vaccination included disbelief in COVID-19 misinformation and disagreement with vaccine conspiracies. Higher COVID-19 vaccine uptake was significantly associated with history of COVID-19 infection, higher income, residence outside the capital, disbelief in COVID-19 misinformation, disagreement with vaccine conspiracies, and reliance on reputable information sources. COVID-19 vaccine coverage prevailed among the participants, with a majority having positive attitudes towards COVID-19 vaccination. Disbelief in COVID- 19 misinformation and disagreement with vaccine conspiracies were correlated with positive vaccine attitudes and higher vaccine uptake. These insights can inform targeted interventions to enhance vaccination campaigns.

## Introduction

Vaccination represents one of the greatest achievements of modern science, with resounding success in controlling the morbidity and mortality associated with infectious diseases.^1-3^ The remarkable success that accompanied the advent of several effective and safe vaccines is manifested by the eradication of smallpox and the control of diseases such as measles and poliomyelitis.^4^

Despite the role of vaccination as a central measure in infectious disease prevention, vaccine hesitancy emerged as a threatening challenge undermining the success of vaccination.^5^ Vaccine hesitancy, defined as the reluctance or rejection of vaccines despite their availability, has become a top threatening global health concern.^6-8^ The issue of vaccine hesitancy emerged long before the emergence of the novel severe acute respiratory syndrome coronavirus 2 (SARS-CoV-2) and the subsequent coronavirus disease 2019 (COVID-19) pandemic.^9-11^

Previous studies indicated a wide range of factors linked with vaccination hesitancy, which is a place-, time-, and context-specific phenomenon.^6, 12-14^ Nevertheless, the emergence of the COVID-19 pandemic exacerbated the issue of vaccine hesitancy.^15,16^ Unsubstantiated conspiracy theories, myths, and mis-/dis-information surrounding the virus, preventive measures, and COVID-19 vaccines fueled the phenomenon of COVID-19 vaccine hesitancy, which was reported in various regions worldwide.^16-18^ Thus, effective control of COVID-19 relies not only on the availability of effective and safe vaccines, but extends to involve positive attitudes and behaviors towards vaccination as well.^19^

The COVID-19 pandemic resulted in a profound negative impact on varying levels. The development and distribution of vaccines emerged as a promising measure to mitigate the negative impact of this unprecedented pandemic.^20, 21^ However, and as mentioned earlier, the success of COVID-19 vaccination campaigns extended beyond the issues of COVID-19 vaccine efficacy and safety, since attitudes towards COVID-19 vaccination played a major role in its uptake.^22^

The infiltration of conspiracy beliefs and misinformation regarding various infectious diseases and vaccines —including COVID-19— has recently been notable in Arab countries.^23-25^ This included unsubstantiated claims which lacked credible scientific evidence. Examples of these false ideas include the idea that SARS-CoV-2 is a man-made virus, the claim that COVID-19 vaccination aimed to implant microchips for surveillance, and the idea that vaccination induces infertility to reduce the global population size.^23, 26, 27^ The association between these factors and adverse health behaviors has been documented in various settings, including the recurring pattern of a link between vaccine hesitancy and endorsement of these beliefs.^27-29^

Iraq, a Middle Eastern country, has a diverse population of over 41 million in 2021, and the country serves as a distinctive case study of a versatile society.^30^ As of 18 October 2023, Iraq reported 2,465,545 cumulative COVID-19 cases and 25,375 cumulative deaths. COVID-19 vaccination in Iraq started on 2 March 2021, with 19,557,364 vaccine doses administered, benefiting 11,332,925 individuals with at least one dose and 7,944,775 individuals with a complete primary vaccination series as of 18 October 2023.^31^ Four vaccine types —Pfizer/BioNTech, Oxford/AstraZeneca, Sinopharm, and Sputnik V— received approval for use in the country.^32^ Several early studies from Iraq showed varying attitudes towards COVID-19 vaccination and its associated determinants; nevertheless, the majority of these studies did not focus on the role of misinformation and vaccine conspiracies on COVID-19 vaccine uptake.^33-40^

Therefore, the objectives of the current study included the elucidation of possible factors that could be associated with higher COVID-19 vaccine uptake and positive attitudes towards the vaccine among the adult Iraqi population. Specifically, we aimed to assess the possible role of vaccine conspiracy beliefs and COVID-19 misinformation in shaping vaccination attitudes and behaviors. Given the scope of this issue, we aspire to provide novel clues into the role of medical-specific conspiracies on adverse health behaviors, potentially offering valuable contributions to collective attempts for successful vaccination campaigns.

## Materials and Methods

### Study design

This study employed a cross-sectional design, utilizing a self-administered online questionnaire as the data collection tool. Inclusion criteria were specified in the introductory section of the e-survey and encompassed the following: participants had to be Iraqi citizens, possess proficiency in the Arabic language, and be aged 18 years or older.

The survey distribution occurred during 5 August 2022–14 September 2022. The questionnaire was hosted in Google Forms, and the survey language was Arabic. The survey distribution was based on chain-referral sampling starting with the contacts of the authors from Iraq (N.K. and M.A.) using e-mails, the direct messaging application WhatsApp, and social media platforms (Facebook and Twitter). The participants were asked to share the survey with their contacts as well. The survey was anonymous, and no incentives were offered for participation. For those who consented to participate, response to all items was mandatory to eliminate item non-response bias.

The minimum sample size in the current study was estimated at 2401 participants. Calculation of the minimum sample size in this study was done using Epitools— Epidemiological Calculators, using the following assumptions: an estimated prevalence of 50%, the desired precision of estimate at 2%, and the Iraqi population size of about 41,179,351 people in 2021.^30, 41^

### Survey instrument

The survey instrument comprised seven sections as follows: First, an introductory section which provided a short explanation of the research and its aims stressing on the anonymity of the survey and the eligibility criteria, followed by the e-consent mandatory item: “Do you agree to participate in the study?”. If the respondent selected “No”, the survey was directed to “submit form” page.

Second, the socio-demographics section: Seven items were used to assess the socio- demographic characteristics of the participants as follows: (1) age (as a continuous variable, and grouped during the analysis based on the median age of the participants into ≤ 27 years vs. > 27 years); (2) sex (male vs. female); (3) occupational category (employed healthcare worker (HCW), employed non-HCW, unemployed, or university/college student); (4) governorate (the Capital Baghdad vs. outside the Capital); (5) educational level (high school or less vs. undergraduate vs. postgraduate); (6) monthly income of household (≤ 500,000 Iraqi dinar (IQD) vs. > 500,000 IQD). The classification was based on two times estimate of the minimum wage in the country (500,000 IQD ≅ 342.55529 US Dollars;^42, 43^ and (7) history of chronic diseases (e.g., diabetes mellitus, hypertension, heart, or renal disease) with yes vs. no as the possible answers.

Third, a section on COVID-19 history of infection and COVID-19 vaccine uptake: Five items were used as follows: (1) “Have you been diagnosed with COVID-19?” (yes vs. no); (2) for the respondents who answered the previous question with “yes”, the question “How many times have you been diagnosed with COVID-19?” followed; (3) “Have you received COVID-19 vaccination” (yes vs. no); (4) for the respondents who answered the previous question with “yes”, the question “Which type of COVID-19 vaccines have you received?” followed with the following choices: the US-based Pfizer-BioNTech COVID-19 Vaccine, the UK-based Oxford–AstraZeneca COVID-19 vaccine, the China-based Sinopharm BBIBP vaccine, the Russia-based Sputnik V vaccine, other types of vaccines and more than one type of vaccine; and (5) “How many doses of COVID-19 vaccines have you received?”

Fourth, a section on the attitude of the participants towards COVID-19 vaccination, comprised the following question: “In your personal opinion, how would you rate the importance of getting the vaccine to protect against COVID-19?”. The item was measured on a 5-point Likert scale: very important, important, neutral/no opinion, not important, and not important at all. These responses were subsequently categorized into three distinct vaccination attitude groups as follows: positive attitude group comprising participants who indicated that COVID-19 vaccination was either “very important” or “important”, the neutral attitude group encompassing participants who answered “neutral/no opinion”, and the negative attitude group including participants who perceived COVID-19 vaccination as “not important” or “not important at all”.

Fifth, assessment of COVID-19 misinformation, with three items based on previous studies addressing COVID-19 vaccine hesitancy in the Middle East,^27, 44^ as follows: (1) “What is your belief regarding the origin of SARS-CoV-2 in humans?” with options to choose “natural” (scored as 1) or “man-made” (scored as 2).; (2) “COVID-19 vaccine is a way to implant microchips into people as a control scheme” (with “yes” scored as 2 vs. “no” scored as 1); (3) “Getting COVID-19 vaccines will lead to infertility” (with “yes” scored as 2 vs. “no” scored as 1). The sum of the scores for the three items comprised the misinformation score which was subsequently classified into four categories as follows: a score of 3 indicated absence of belief in COVID-19 misinformation, a score of 4 indicated a slight belief in misinformation, a score of 5 indicated a moderate belief in misinformation, and a score of 6 indicating a strong belief in COVID-19 misinformation.

Sixth, assessment of the sources of COVID-19 vaccine information using the following item: “What is your single main source of information about COVID-19 vaccines?” with the following options: Physicians, scientists and scientific journals vs. TV programs and newspapers vs. social media platforms.

Finally, the seventh section on the assessment of COVID-19 vaccine conspiracy beliefs: The conspiracy beliefs regarding COVID-19 vaccination was assessed using seven items based on the original vaccine conspiracy beliefs scale (VCBS) adopted from Shapiro *et al*. that was used previously in the assessment of COVID-19 vaccine hesitancy and influenza vaccine uptake.^27, 45, 46^ The seven items were: (1) COVID-19 vaccine safety data are often fabricated; (2) people are deceived about the safety of COVID-19 vaccines (3) pharmaceutical companies cover up the dangers of COVID-19 vaccines; (4) COVID-19 vaccine efficacy data are often fabricated; (5) people are deceived about the effectiveness of COVID-19 vaccines; (6) vaccination of children is harmful, and this fact is hidden from people; and (7) governments are trying to cover up the link between vaccines and other diseases, such as autism.^27, 45^ The scale was assessed on a 7-point Likert scale as follows: strongly disagree (scored as 1), disagree (scored as 2), somewhat disagree (scored as 3), neutral/no opinion (scored as 4), somewhat agree (scored as 5), agree (scored as 6) and strongly agree (scored as 7). The Cronbach α value for the VCBS was 0.893 indicating the acceptable internal consistency of the scale. The VCBS score could range between 7 and 49, with higher scores indicating the embrace of vaccine conspiracy beliefs. The VCBS was then classified into three categories as follows: VCBS: 7–20 (disagreement with vaccine conspiracies), VCBS: 21–35 (neutral), and VCBS: 36–49 (agreement with vaccine conspiracies).

### Ethical approval

The current study was approved by the Scientific and Ethical Unit at Al-Kindy College of Medicine, University of Baghdad (approved by the Council of Al-Kindy College of Medicine in session No. 20, date: 6 July 2022 under no specific reference number).

An electronic informed consent was required for successful completion of the survey by the inclusion of an introductory mandatory item asking the participants to approve their participation in the survey.

### Statistical analysis

Data and statistical analyses were conducted using BM SPSS v26.0 for Windows (Armonk, NY, USA: IBM Corp.). Univariate analyses were conducted employing the chi-squared (χ^2^) test. Associations with a significance level of *p*<.100 in univariate analyses were considered for inclusion in the subsequent multivariate analysis. Multivariate analysis was conducted using multinomial logistic regression. The Nagelkerke R^2^ statistic was employed to assess the variance explained by the model. A significance threshold of *p*<.050 was applied to determine statistical significance.

## Results

### Study sample characteristics

The study sample comprised a total of 2544 individuals. Characteristics of the study sample are shown in (**Table 1**).

**Table 1.**
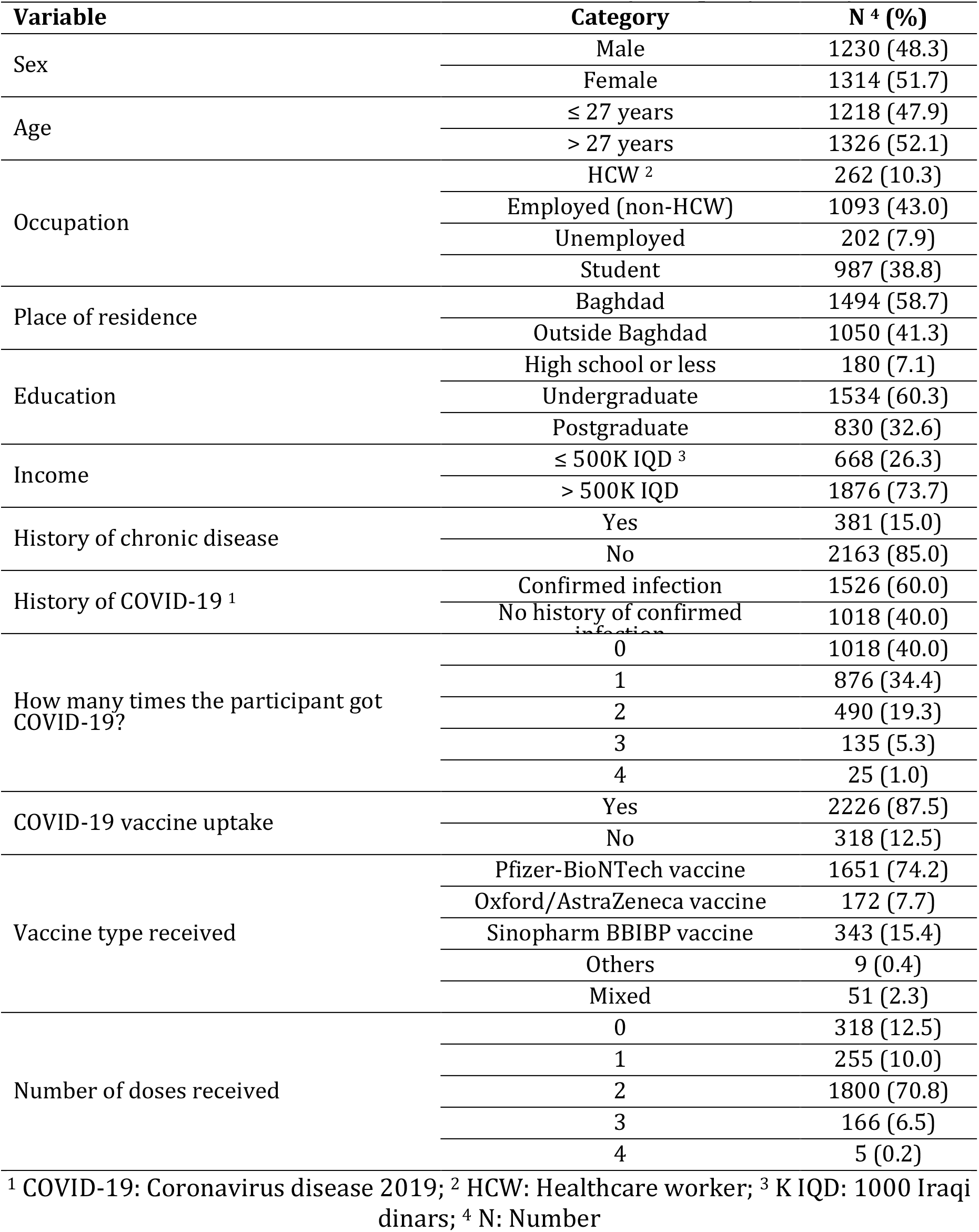
General characteristics of the study sample (*N*=2544).

### Attitude Towards COVID-19 Vaccination, COVID-19 Vaccine Uptake and its Associated Variables

The overall attitude of the participants towards COVID-19 vaccination was mostly positive (n=1966, 77.3%), while a neutral attitude was expressed by 345 participants (13.6%), and negative attitude was expressed by 233 participants (9.2%).

Using univariate analysis, the following factors were associated with a positive attitude towards COVID-19 vaccination: male sex, age > 27 years, being a HCW, postgraduate education, and monthly income > 500K IQD (**Table 2**).

**Table 2.** Variables associated with attitude to COVID-19 vaccination and COVID-19 vaccine uptake in univariate analysis.

| Variable | Category | Attitude towards COVID-19 vaccination | | | p, $\chi^2$ | Self-reported history of COVID-19 vaccine uptake | | p, $\chi^2$ |
| --- | --- | --- | --- | --- | --- | --- | --- | --- |
|  |  | Positive N<sup>4</sup> (%) | Neutral N (%) | Negative N (%) |  | Yes N (%) | No N (%) |  |
| Sex | Male | 991 (80.6) | 124 (10.1) | 115 (9.3) | <.001, 24.695 | 1092 (88.8) | 138 (11.2) | .059, 3.570 |
|  | Female | 975 (74.2) | 221 (16.8) | 118 (9.0) |  | 1134 (86.3) | 180 (13.7) |  |
| Age | ≤ 27 years | 898 (73.7) | 206 (16.9) | 114 (9.4) | <.001, 23.276 | 1069 (87.8) | 149 (12.2) | .697, 0.152 |
|  | > 27 years | 1068 (80.5) | 139 (10.5) | 119 (9.0) |  | 1157 (87.3) | 169 (12.7) |  |
| Occupation | HCW<sup>2</sup> | 222 (84.7) | 27 (10.3) | 13 (5.0) | <.001, 24.264 | 248 (94.7) | 14 (5.3) | .002, 14.561 |
|  | Employed (non-HCW) | 869 (79.5) | 123 (11.3) | 101 (9.2) |  | 940 (86.0) | 153 (14.0) |  |
|  | Unemployed | 151 (74.8) | 32 (15.8) | 19 (9.4) |  | 176 (87.1) | 26 (12.9) |  |
|  | Student | 724 (73.4) | 163 (16.5) | 100 (10.1) |  | 862 (87.3) | 125 (12.7) |  |
| Place of residence | Baghdad | 1141 (76.4) | 214 (14.3) | 139 (9.3) | .364, 2.022 | 1274 (85.3) | 220 (14.7) | <.001, 16.392 |
|  | Outside Baghdad | 825 (78.6) | 131 (12.5) | 94 (9.0) |  | 952 (90.7) | 98 (9.3) |  |
| Education | High school or less | 133 (73.9) | 30 (16.7) | 17 (9.4) | <.001, 25.358 | 149 (82.8) | 31 (17.2) | .135, 4.002 |
|  | Undergraduate | 1145 (74.6) | 240 (15.6) | 149 (9.7) |  | 1346 (87.7) | 188 (12.3) |  |
|  | Postgraduate | 688 (82.9) | 75 (9.0) | 67 (8.1) |  | 731 (88.1) | 99 (11.9) |  |
| Income | ≤ 500K IQD<sup>3</sup> | 481 (72.0) | 114 (17.1) | 73 (10.9) | .001, 14.561 | 557 (83.4) | 111 (16.6) | <.001, 14.036 |
|  | > 500K IQD | 1485 (79.2) | 231 (12.3) | 160 (8.5) |  | 1669 (89.0) | 207 (11.0) |  |
| History of chronic disease | Yes | 292 (76.6) | 54 (14.2) | 35 (9.2) | .929, 0.148 | 340 (89.2) | 41 (10.8) | .266, 1.239 |
|  | No | 1674 (77.4) | 291 (13.5) | 198 (9.2) |  | 1886 (87.2) | 277 (12.8) |  |
| History of COVID-19<sup>1</sup> | Confirmed infection | 1171 (76.7) | 218 (14.3) | 137 (9.0) | .415, 1.758 | 1362 (89.3) | 164 (10.7) | .001, 10.714 |
|  | No history of confirmed infection | 795 (78.1) | 127 (12.5) | 96 (9.4) |  | 864 (84.9) | 154 (15.1) |  |

<sup>1</sup> COVID-19: Coronavirus disease 2019; <sup>2</sup> HCW: Healthcare worker; <sup>3</sup> K IQD: 1000 Iraqi dinars; <sup>4</sup> N: Number

The majority of participants reported uptake of at least a single dose of COVID-19 vaccination (n=2226, 87.5%) while 318 participants had no self-reported history of COVID-19 vaccine uptake (12.5%). Using univariate analysis, the following factors were associated with COVID-19 vaccine uptake: being an HCW, residence outside the Capital, income > 500K IQD, and a history of confirmed COVID-19 infection (**Table 2**).

### The Belief in COVID-19 Misinformation and the Embrace of COVID-19 Vaccine Conspiracy Beliefs

Regarding the belief in COVID-19 misinformation, the complete absence of such beliefs was reported among 731 participants (28.7%), while a slight belief in misinformation was reported among 922 participants (36.2%). Moderate belief in misinformation was reported among 576 participants (22.6%), and the strong belief in COVID-19 misinformation was observed among 315 participants (12.4%). For the attitude towards COVID-19 vaccine conspiracies, the majority of participants exhibited a neutral attitude (n=1464, 57.5%), with 607 participants showing the embrace of COVID-19 vaccine conspiracies (23.9%), and 473 showing disagreement with such beliefs (18.6%).

In univariate analysis, the strong belief in COVID-19 misinformation and the agreement with COVID-19 vaccine conspiracies were associated with both negative attitude to COVID-19 vaccination and less COVID-19 vaccine uptake (**Table 3**).

**Table 3.** Association between attitude to vaccine conspiracies, COVID-19 misinformation and attitude to COVID-19 vaccination/COVID-19 vaccination uptake in univariate analysis.

| Variable | Category | Attitude towards COVID-19 vaccination | | | p, $\chi^2$ | COVID-19 vaccine uptake | | p, $\chi^2$ |
| --- | --- | --- | --- | --- | --- | --- | --- | --- |
|  |  | Positive N<sup>4</sup> (%) | Neutral N (%) | Negative N (%) |  | Yes N (%) | No N (%) |  |
| Misinformation score<sup>1</sup> | No belief in misinformation | 665 (91.0) | 51 (7.0) | 15 (2.1) | <.001, 303.899 | 688 (94.1) | 43 (5.9) | <.001, 75.253 |
|  | Slight belief in misinformation | 767 (83.2) | 102 (11.1) | 53 (5.7) |  | 820 (88.9) | 102 (11.1) |  |
|  | Moderate belief in misinformation | 381 (66.1) | 110 (19.1) | 85 (14.8) |  | 475 (82.5) | 101 (17.5) |  |
|  | Strong belief in misinformation | 153 (48.6) | 82 (26) | 80 (25.4) |  | 243 (77.1) | 72 (22.9) |  |
| Attitude towards COVID-19 vaccine conspiracy<sup>2</sup> | Disagreement (VCBS<sup>3</sup>: 7–20) | 455 (96.2) | 11 (2.3) | 7 (1.5) | <.001, 346.968 | 452 (95.6) | 21 (4.4) | <.001, 96.532 |
|  | Neutral (VCBS: 21–35) | 1168 (79.8) | 224 (15.3) | 72 (4.9) |  | 1308 (89.3) | 156 (10.7) |  |
|  | Agreement (VCBS: 36–49) | 343 (56.5) | 110 (18.1) | 154 (25.4) |  | 466 (76.8) | 141 (23.2) |  |

<sup>1</sup> Misinformation score: Using three items to assess the belief in COVID-19 misinformation; <sup>2</sup> COVID-19: Coronavirus disease 2019; <sup>3</sup> VCBS: Vaccine conspiracy beliefs scale; <sup>4</sup> N: Number.

### Source of Information about COVID-19 Vaccines

The main sources of information regarding COVID-19 vaccination included physicians, scientists, and scientific journals (n=1090, 42.8%), followed closely by social media platforms (n=1040, 40.9%), and finally TV programs and newspapers (n=414, 16.3%). The dependence on social media platforms was associated with both a negative attitude to COVID-19 vaccination and less COVID-19 vaccine uptake (**Figure 1**).

**Figure 1:**
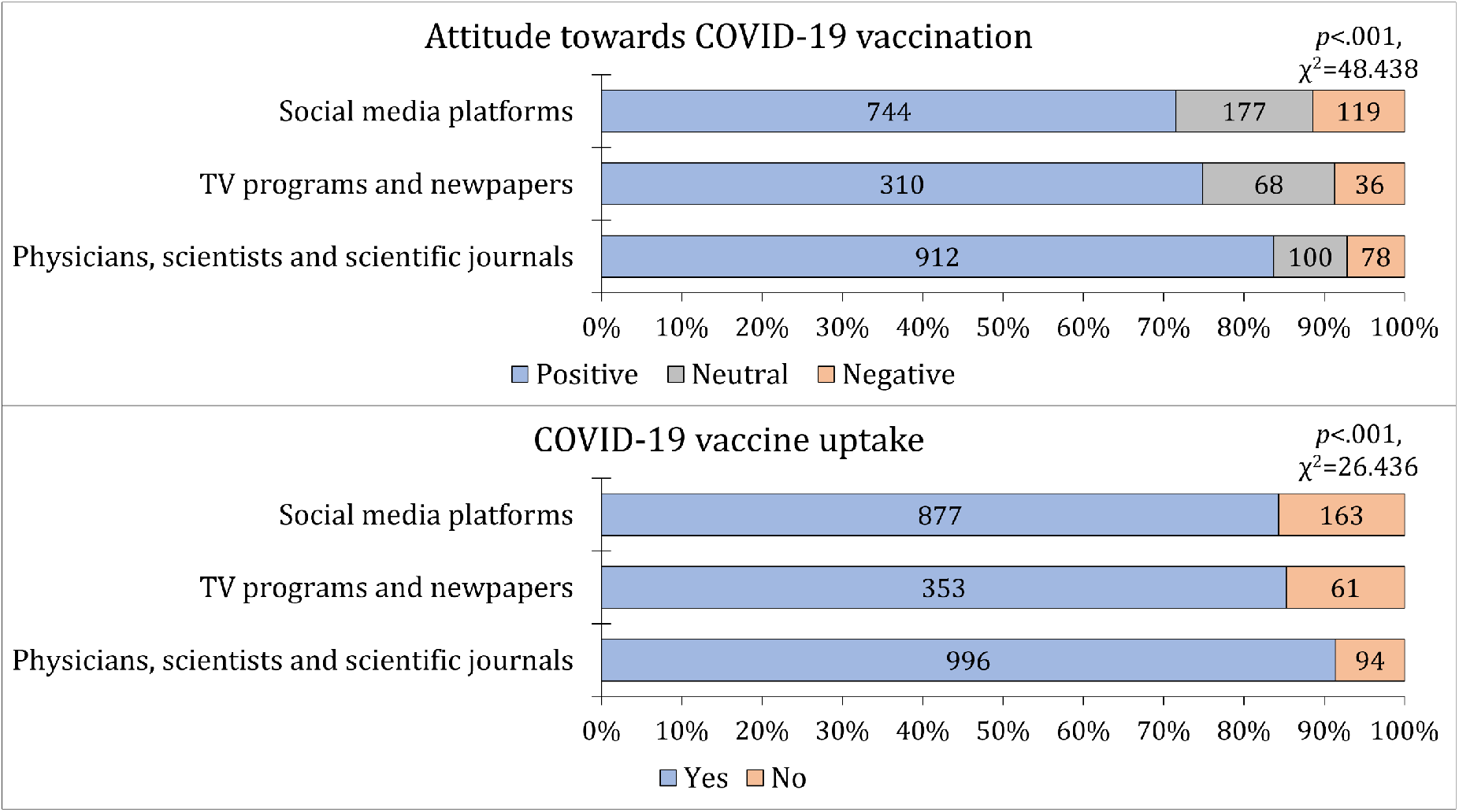
The association between attitude towards COVID-19 vaccination, COVID-19 vaccine uptake and the main source of information regarding the vaccine. COVID-19: Coronavirus disease 2019.

### Multivariate analysis for factors associated with Positive Attitude towards COVID- 19 Vaccination

The Nagelkerke R^2^ value of 0.245 indicated that the regression model explained 24.5% of the variability observed in the data. For demographic variables, only males were significantly less likely to have a neutral attitude compared to females, with an adjusted odds ratio (aOR) of 0.64 (95% CI: 0.44–0.92, *p*=.016).

Statistically significant associations were observed between belief in misinformation and COVID-19 vaccine attitude. Participants with no belief in misinformation displayed a significantly higher likelihood of a positive attitude, with an aOR of 7.82 (95% CI: 4.16– 14.68), while those with slight belief exhibited an aOR of 3.75 (95% CI: 2.46–5.71), and those with moderate belief an aOR of 1.58 (95% CI: 1.07–2.31), all compared to strong belief (*p*<.001, **Table 4**). Similarly, those who disagreed with COVID-19 vaccine conspiracy beliefs (VCBS: 7–20) showed a higher likelihood of a positive attitude towards COVID-19 vaccination, with an aOR of 10.42 (95% CI: 4.62–23.54), while those with a neutral attitude (VCBS: 21–35) displayed an aOR of 4.57 (95% CI: 3.30–6.33), both compared to those who endorsed vaccine conspiracies (*p*<.001, **Table 4**). Additionally, participants with a neutral vaccine conspiracy attitude (VCBS: 21–35) displayed a higher likelihood of a neutral attitude towards COVID-19 vaccination, with an aOR of 4.02 (95% CI: 2.73–5.92), compared to those who endorsed vaccine conspiracies (*p*<.001, **Table 4**).

**Table 4.** Variables associated with attitude to COVID-19 vaccination in multinomial logistic regression analyses.

| Model | Nagelkerke R<sup>2</sup> = .245 | aOR <sup>6</sup> (95% CI <sup>7</sup>) | p value |
| --- | --- | --- | --- |
| **Positive attitude vs. negative attitude towards COVID-19 vaccination** |  |  |  |
| Sex | Male | 0.99 (0.73–1.35) | .951 |
|  | Female | Ref. | . |
| Age | ≤ 27 years | 0.93 (0.60–1.45) | .763 |
|  | > 27 years | Ref. | . |
| Occupation | HCW <sup>3</sup> | 1.52 (0.77–2.98) | .229 |
|  | Employed (non-HCW) | 1.04 (0.67–1.63) | .857 |
|  | Unemployed | 1.03 (0.58–1.83) | .922 |
|  | Student | Ref. | . |
| Education | High school or less | 0.80 (0.42–1.54) | .506 |
|  | Undergraduate | 0.74 (0.50–1.10) | .139 |
|  | Postgraduate | Ref. | . |
| Income | ≤ 500K IQD <sup>4</sup> | 1.03 (0.73–1.46) | .873 |
|  | > 500K IQD | Ref. | . |
| Misinformation score <sup>1</sup> | No belief in misinformation | 7.82 (4.16–14.68) | <.001 |
|  | Slight belief in misinformation | 3.75 (2.46–5.71) | <.001 |
|  | Moderate belief in misinformation | 1.58 (1.07–2.31) | .020 |
|  | Strong belief in misinformation | Ref. | . |
| Attitude towards COVID-19 <sup>2</sup> vaccine conspiracy | Disagreement (VCBS <sup>5</sup>: 7–20) | 10.42 (4.62–23.54) | <.001 |
|  | Neutral (VCBS: 21–35) | 4.57 (3.30–6.33) | <.001 |
|  | Agreement (VCBS: 36–49) | Ref. | . |
| Source of information regarding COVID-19 vaccination | Physicians, scientists and scientific journals | 1.26 (0.90–1.75) | .179 |
|  | TV programs and newspapers | 1.43 (0.93–2.19) | .105 |
|  | Social media platforms | Ref. | . |
| **Neutral attitude vs. negative attitude towards COVID-19 vaccination** |  |  |  |
| Sex | Male | 0.64 (0.44–0.92) | .016 |
|  | Female | Ref. | . |
| Age | ≤ 27 years | 1.38 (0.83–2.31) | .217 |
|  | > 27 years | Ref. | . |
| Occupation | HCW | 1.49 (0.69–3.21) | .314 |
|  | Employed (non-HCW) | 1.17 (0.69–1.96) | .562 |
|  | Unemployed | 1.10 (0.57–2.11) | .774 |
|  | Student | Ref. | . |
| Education | High school or less | 1.31 (0.61–2.78) | .488 |
|  | Undergraduate | 1.17 (0.73–1.87) | .512 |
|  | Postgraduate | Ref. | . |
| Income | ≤ 500K IQD <sup>3</sup> | 1.00 (0.68–1.49) | .987 |
|  | > 500K IQD | Ref. | . |
| Misinformation score | No belief in misinformation | 1.74 (0.85–3.54) | .128 |
|  | Slight belief in misinformation | 1.09 (0.67–1.78) | .730 |
|  | Moderate belief in misinformation | 0.89 (0.57–1.39) | .613 |
|  | Strong belief in misinformation | Ref. | . |
| Attitude towards COVID-19 vaccine conspiracy | Disagreement (VCBS <sup>3</sup>: 7–20) | 1.71 (0.61–4.79) | .306 |
|  | Neutral (VCBS: 21–35) | 4.02 (2.73–5.92) | <.001 |
|  | Agreement (VCBS: 36–49) | Ref. | . |
| Source of information regarding COVID-19 vaccination | Physicians, scientists and scientific journals | 0.85 (0.57–1.26) | .415 |
|  | TV programs and newspapers | 1.38 (0.85–2.25) | .191 |
|  | Social media platforms | Ref. | . |

<sup>1</sup> Misinformation score: Using three items to assess the belief in COVID-19 misinformation; <sup>2</sup> COVID-19: Coronavirus disease 2019; <sup>3</sup> HCW: Healthcare worker; <sup>4</sup> K IQD: 1000 Iraqi dinars; <sup>5</sup> VCBS: Vaccine conspiracy beliefs scale; <sup>6</sup> aOR: Adjusted odds ratio; <sup>7</sup> CI: Confidence interval.

### Multivariate Analysis for the Factors Associated with COVID-19 Vaccine Uptake

The Nagelkerke R^2^ of 0.128 showed that the regression model explained 12.8% of the variability observed in the data. Participants residing in Baghdad were less likely to have received the COVID-19 vaccine, as indicated by an aOR of 0.56 (95% CI: 0.43–0.73, *p*<.001, **Table 5**), compared to those residing outside Baghdad. Employed individuals in non-healthcare roles were significantly less likely to have received the COVID-19 vaccine, with an aOR of 0.70 (95% CI: 0.52–0.93, *p*=.015, **Table 5**) compared to university/college students. Individuals with a history of COVID-19 infection showed higher rates of COVID- 19 vaccine uptake, with an aOR of 1.53 (95% CI: 1.19–1.97, *p*=.001, **Table 5**). Additionally, participants with an income of ≤ 500K IQD showed less likelihood of COVID-19 vaccine uptake, with an aOR of 0.66 (95% CI: 0.50–0.88), compared to those with incomes exceeding 500K IQD (*p*=.004, **Table 5**).

**Table 5.** Variables associated with COVID-19 vaccine uptake in multinomial logistic regression analyses.

| Model | Nagelkerke $R^2 = .128$ | aOR <sup>6</sup> (95% CI <sup>7</sup>) | $p$ value |
| --- | --- | --- | --- |
| **COVID-19 vaccine uptake vs. no history of COVID-19 vaccination** |  |  |  |
| Sex | Male | 1.27 (0.97–1.64) | .079 |
|  | Female | Ref. | . |
| Place of residence | Baghdad | 0.56 (0.43–0.73) | **<.001** |
|  | Outside Baghdad | Ref. | . |
| Occupation | HCW <sup>3</sup> | 1.69 (0.93–3.07) | 0.083 |
|  | Employed (non-HCW) | 0.70 (0.52–0.93) | **0.015** |
|  | Unemployed | 1.00 (0.62–1.60) | 0.985 |
|  | Student | Ref. | . |
| History of COVID-19 | Yes | 1.53 (1.19–1.97) | **0.001** |
|  | No | Ref. | . |
| Income | $\leq 500K$ IQD <sup>4</sup> | 0.66 (0.50–0.88) | **0.004** |
| | $> 500K$ IQD | Ref. | . |
| Misinformation score <sup>1</sup> | No belief in misinformation | 2.29 (1.45–3.61) | **<.001** |
|  | Slight belief in misinformation | 1.49 (1.04–2.15) | **0.031** |
|  | Moderate belief in misinformation | 1.08 (0.75–1.54) | 0.687 |
|  | Strong belief in misinformation | Ref. | . |
| Attitude towards COVID-19 <sup>2</sup> vaccine conspiracy | Disagreement (VCBS <sup>5</sup>: 7–20) | 3.65 (2.16–6.18) | **<.001** |
|  | Neutral (VCBS: 21–35) | 2.06 (1.56–2.71) | **<.001** |
|  | Agreement (VCBS: 36–49) | Ref. | . |
| Source of information regarding COVID-19 vaccination | Physicians, scientists and scientific journals | 1.46 (1.10–1.94) | **0.009** |
|  | TV programs and newspapers | 1.13 (0.81–1.58) | 0.469 |
|  | Social media platforms | Ref. | . |

<sup>1</sup> Misinformation score: Using three items to assess the belief in COVID-19 misinformation; <sup>2</sup> COVID-19: Coronavirus disease 2019; <sup>3</sup> HCW: Healthcare worker; <sup>4</sup> K IQD: 1000 Iraqi dinars; <sup>5</sup> VCBS: Vaccine conspiracy beliefs scale; <sup>6</sup> aOR: Adjusted odds ratio; <sup>7</sup> CI: Confidence interval.

Moreover, participants who reported no belief in COVID-19 misinformation exhibited a significantly higher likelihood of COVID-19 vaccine uptake, reflected in an aOR of 2.29 (95% CI: 1.45–3.61) compared to those with strong beliefs in misinformation (*p*<.001). Also, participants with a slight belief in misinformation demonstrated a higher likelihood of COVID-19 vaccine uptake, with an aOR of 1.49 (95% CI: 1.04–2.15, *p*=.031, **Table 5**). A higher likelihood of COVID-19 vaccine uptake was observed among the participants who disagreed with COVID-19 vaccine conspiracy beliefs (VCBS: 7–20) reflected by an aOR of 3.65 (95% CI: 2.16–6.18), and among participants with neutral attitude towards vaccine conspiracies (VCBS: 21–35) with an aOR of 2.06 (95% CI: 1.56–2.71) compared to those who endorsed vaccine conspiracies (*p*<.001 for both comparisons, **Table 5**). Finally, participants who relied on physicians, scientists, and scientific journals as their source of information regarding COVID-19 vaccination exhibited a significantly increased likelihood of COVID-19 vaccine uptake, as indicated by an aOR of 1.46 (95% CI: 1.10–1.94, *p*=.009, **Table 5**) compared to those who relied on social media platforms.

## Discussion

The current study revealed a clear distinct pattern in the possible factors influencing COVID-19 vaccination attitudes and uptake. Notably, lower vaccine uptake and negative attitudes towards the vaccine were significantly associated with endorsement of vaccine conspiracies and COVID-19 misinformation.

In comparison to the previous studies conducted in Iraq, the participants in the current study demonstrated a favorable attitude towards COVID-19 vaccination. For example, an earlier study by Ghazi *et al*. indicated that 77.6% of respondents were willing to take the vaccine when available, a rate almost the same of the current study findings which indicated that 77.3% displayed a positive attitude to COVID-19 vaccination.^33^ Another Iraqi study in 2021 by Shareef *et al*. reported a lower acceptance rate of 56.2%.^37^ On the other hand, Al-Qerem *et al*. conducted a survey study in July 2021 and found that 88.6% of respondents were willing to be vaccinated against COVID-19, with concerns about vaccine safety and the need for more information being the primary reasons for vaccine refusal.^38^

From a global perspective, the acceptance rate observed in our study is slightly higher than the global average of 65–75%, as indicated by recent systematic reviews.^47, 48^ Despite the observed variability in the rates of COVID-19 vaccine acceptance which can be attributed to survey timing, phrasing of the items assessing vaccination hesitancy, and possible sampling bias, the common pattern in line with our findings is the generally positive attitude towards COVID-19 vaccination in Iraq.^16, 37-39^

A notable aspect of this study was the clear demonstration of the correlation between COVID-19 vaccine conspiracies, COVID-19 misinformation, and negative attitudes and behaviors towards COVID-19 vaccination. This manifested in significantly lower vaccine uptake and less favorable attitudes towards the vaccine. The government in Iraq has taken measures to combat such misinformation, emphasizing the importance of vaccination and warning against spreading false information.^49, 50^

The Iraqi government adopted a non-mandatory approach regarding COVID-19 vaccination, refraining from imposing vaccine mandates due to the lack of legal support of such a measure.^51^ Instead, the Iraqi Ministry of Health and Environment advocated for alternative public health strategies. In Iraq, employees were encouraged to voluntarily provide either a weekly COVID-19 testing card or a vaccination card as a prerequisite for work attendance without punishing measures against unvaccinated employees.^51^ The Iraqi government also initiated vaccination campaigns to enhance COVID-19 vaccine coverage, with comprehensive plans to augment vaccine supply and to expand vaccination facilities, with a particular focus on vaccinating HCWs to counter vaccine-related misinformation effectively.^51, 52^

The findings of the current study highlighted the significant association between conspiracy beliefs and negative health behavior manifested in lower COVID-19 vaccine uptake. Extensive evidence has consistently highlighted the widespread presence of medical conspiracy theories and their potential impact on various aspects of health-related behaviors, including the willingness to get vaccinated and actual vaccine uptake.^27, 46, 53-157^ The adoption of conspiracy theories can exert a direct influence on individual engagement behaviors, including health-related practices.^58-60^ For example, the detrimental impact of embracing COVID-19 conspiracy beliefs on compliance with government-imposed restrictions, adherence to preventive measures, and willingness to receive COVID-19 vaccination was shown in a study from the U.S. by Romer and Jamieson.^61^ Similarly, research conducted in Finland by Pivetti *et al*. demonstrated that the endorsement of COVID-19 conspiracy beliefs was associated with lower support for pandemic-related governmental restrictions.^62^ In the Arab countries of the Middle East, the COVID-19 vaccine conspiracies have been shown to be associated with higher rates of vaccine hesitancy/rejection.^27, 44^

In the current study, an interesting observation was the striking contrast in the determinants of participants’ attitudes towards COVID-19 vaccination compared to their actual vaccine uptake. Notably, demographic variables appeared to play a minimal role in shaping attitudes, whereas several demographic variables were significantly correlated with actual vaccine uptake. This divergence between attitudes and behavior can be attributed to the inherent distinction between what people think or feel representing attitudes, and what they actually do manifested in behavior.^63^ Attitudes often reflect abstract viewpoints and personal beliefs, while behavior could be influenced by a range of external factors, including governmental policies and societal expectations.^64, 65^ Thus, it is conceivable that individuals may hold certain attitudes about vaccination but, when faced with practical circumstances, their behavior may align differently.

The major finding in this study was unveiling the significant association between vaccine conspiracy beliefs, misinformation, and negative attitudes towards COVID-19 vaccination, as well as a reduced likelihood of vaccine uptake. Plausible explanations of this association could be based on previous and recent evidence highlighting the impact of conspiracies and misinformation on vaccination behavior.^54,66-68^ Conspiracy theories and misinformation have the potential to deter individuals from getting vaccinated through reducing vaccine confidence.^18, 69^ Endorsing conspiracy beliefs can undermine the trust in healthcare systems, governmental agencies, and pharmaceutical companies.^70,71^ Trust is an essential aspect in the decision-making process of getting vaccinated.^72, 73^ Hence, compromised trust could result in a negative attitudes towards vaccination due to fear among individuals that they are not being provided with accurate and safe preventive measures.^74^

Additionally, the current study results showed that the source of information could play an important role in COVID-19 vaccine uptake. Health misinformation often spreads through channels that may lack credibility, such as social media platforms.^75-77^ When individuals rely on social media for health information, they may inadvertently expose themselves to distorted views on vaccine safety and efficacy.^78^ The current study results were consistent with this point of view by revealing that COVID-19 vaccine uptake was lower among those who relied on social media platforms compared to individuals who sought vaccine information from scientifically credible sources (e.g., physicians, scientists, etc.).

Besides the important roles of vaccine conspiracy beliefs and misinformation, it is worth mentioning other factors were linked to actual COVID-19 vaccine uptake in this study. These factors could offer useful insights into the complexity of the vaccine uptake as a health behavior. First, participants living outside of the capital, Baghdad exhibited higher COVID-19 vaccine uptake. This suggests that targeted efforts should be made to prioritize vaccination campaigns in the capital city of Iraq. Additionally, lower income was associated with lower COVID-19 vaccine uptake, which emphasizes the importance of addressing socio-economic disparities to ensure equitable access to COVID-19 vaccination. Furthermore, the history of COVID-19 infection appeared to influence vaccination behavior, although the exact nature of this relationship is not discernible. It is possible that individuals who had experienced the disease may have been less complacent about vaccination due to their personal encounter with the disease. However, establishing a direct cause-and-effect relationship in this regard can be challenging and requires further investigation. Finally, in this study, university/college students, as a group, exhibited higher COVID-19 vaccination rates. This aligns with previous research and can be attributed to the view that students are generally more informed and engaged in health-related matters.^44, 79^

Lastly, it is essential to acknowledge the limitations of this study, which should be considered carefully in any attempt to generalize the results as follows: (1) The possibility of response bias should be considered with participants who chose to respond to the survey being not representative of the entire adult population in Iraq. Additionally, individuals with stronger opinions, whether positive or negative, about COVID-19 vaccination might have been more motivated to participate in the study, resulting in response bias as well, with subsequent over- or under-estimation of COVID-19 vaccine hesitancy/resistance and misinformation levels. (2) The study employed a cross-sectional design, entailing the capturing of data at a single point of time, which is helpful for elucidating associations but cannot establish causality. Additionally, the data were collected at a specific time point, making it hard to draw definitive conclusions about the temporal trends in vaccination attitudes and behaviors. It is recommended to conduct longitudinal studies to establish causal relationships and to analyze the temporal trends. (3) The current study utilized the chain-referral sampling method to recruit participants, which is a non-random sampling method possibly introducing selection bias; therefore, the sample may not be fully representative of the broader adult Iraqi population. In addition, the study inevitably excluded certain groups, such as individuals without internet access or those who could not complete an online survey, further deepening the issue of possible selection bias. (4) The findings of this study may not be easily generalizable to other regions or countries with different cultural, social, or healthcare contexts, based on the attributes of vaccine hesitancy as a phenomenon. Subsequently, this could compromise the generalizability of the study results on the global level. (5) Social desirability bias should be considered since the participants might have provided what they believed as socially acceptable responses. This could have led to overreporting positive attitudes towards vaccination and underreporting negative vaccination attitudes endorsing conspiracy beliefs.

## Conclusions

In conclusion, this study aimed to provide a comprehensive assessment of the factors shaping attitudes and behaviors related to COVID-19 vaccination in Iraq. The findings of the current study could provide valuable insights into the interplay between COVID-19 vaccine conspiracies, COVID-19 misinformation, and negative health attitudes and behaviors, which were manifested in lower COVID-19 vaccine uptake and unfavorable attitudes towards the vaccine.

These results emphasized the critical need for targeted interventions aiming to address misinformation and to enhance COVID-19 vaccine literacy. Engaging HCWs as advocates for vaccination can play a pivotal role in improving vaccine acceptance and uptake among the population, which was highlighted by their significant role as a source of information among participants who had higher rates of COVID-19 vaccine uptake. Furthermore, tailoring communication strategies to specific demographics and belief-based subgroups can be an important measure for effectively countering COVID-19 vaccine-related uptake challenges. Ultimately, the implementation of these strategies can contribute to the global effort to combat the COVID-19 pandemic by increasing vaccine acceptance and participation, ultimately resulting in a positive impact on global health.

## Data Availability

The data presented in this study are available on request from the corresponding author (Malik Sallam).

## Acknowledgments

None.

## Supplementary Materials

None.

